# COVID-19 and Mental Health: A Study of its Impact on Students in Maharashtra, India

**DOI:** 10.1101/2020.08.05.20160499

**Authors:** Kshipra Moghe, Disha Kotecha, Manjusha Patil

## Abstract

This study identifies and analyzes the personal, social and psychological impact of COVID - 19 on the mental health of students of age group 16 to 25. A response from N= 351 students provided a comparative analysis based on the gender, and background via t-test with significance factor of p ≤ 0.5, to understand the pandemic’s impact. The results show that female students are more concerned about health, and future, and are more prone to psychological issues like feelings of uncertainty, helplessness and outbursts than male students. Urban student’s population is more mentally affected than their rural counterparts. An increase is seen in need for solitude, being withdrawn and self-harm in male students. A shift in perception from seeing family as a source of support to that of a restriction is indicated, although the benefits of a collectivistic society are undisputed.

**Impact Statement:** This study performs an analysis of the student’s response to questions based on social and self-perception as a result of COVID-19. It also discusses the nature of adaptive strategies espoused by them and their effectiveness in dealing with the pandemic, isolation, and the new normal.

## Introduction

The first documented proof of Novel Coronavirus or COVID-19 was reported in Wuhan city in the Hubei province in China on 31 December 2019. The patients detailed traits of respiratory diseases and pneumonia that required hospitalization (“Pneumonia of unknown cause – China,” 2020). The number of cases began increasing at an alarming pace and a Public Health Emergency of International Concern was declared on 30 January 2020 to tackle this issue. It created a massive uproar while significantly affecting the health of people. In India, Kerala marked the first case of Coronavirus in January 2020 with a patient, who had a travel history from Wuhan. Around 114 countries had been affected by Coronavirus in just two months. Taking account of its escalation intensity and area of influence, on 11 March the WHO declared it a pandemic. Overall, as of 29 June, approximately 216 countries have been affected, with 99, 62, 193 verified cases and 4, 98, 723 affirmed deaths (“Coronavirus Disease (COVID-19) Situation Reports,” n.d.; “Coronavirus Disease (COVID-19) - events as they happen,” n.d.). The virus communicates through minuscule droplets while sneezing, coughing, or contact in close vicinity. Scientists have been working on creating medicine or vaccination for it, but it would take some time (He et al., 2020; Sharma & Nigam, 2020). Therefore, wearing a mask and maintaining a 3ft distance in public is imperative to avoid being infected. To avoid the devastating effect COVID-19 had on Western countries and considering the vast population, India implemented a countywide lockdown on 25 March, with only stores of necessary and basic amenities such as supermarkets and pharmacies allowed to function.

Apart from provoking massive health uproar, this pandemic also seems to have created an economic, mental, and social effect on the masses. The economy of a country and an individual suffer severe damage in situations like these. A reduction in the supply chain creates a scarcity of food, resources, and personal protective equipment (Ebrahim et al., 2020). It leads to a financial strain on society and an imbalance of the economy, especially in a country like India. This expectedly, in turn creates unrest and a general sense of helplessness. Social distancing measures, quarantine, shutting down of educational institutions, and self-isolation have a detrimental impact on people’s psychology due to increased loneliness, distrust, and reduced social interaction. A weak immune system or having closed ones susceptible to diseases intensifies stress, anxiety, and frustration in the populace, as it reaches a life-threatening level. Constant overloading of information called ‘infodemic’ via social media platforms creates uncertainty and worry among the people while risking the spread of false information (Fiorillo & Gorwood, 2020).

Studies have shown that between the age of 16-25, students show stress, anxiety and depressive tendencies (Mahmoud et al., 2012). Although they develop new skills in maintaining relationships, independence and self-sufficiency, any hurdle in this process can cause denial, self-blaming, dissatisfaction, stress or anxiety. Hence, social media works as a coping mechanism with its ease of accessibility and trend, albeit its negative impact on physical and mental health. Specifically, during the lockdown period of COVID-19, factors related to changes in academic structures, examinations and a battle with limited resources can be directly associated with anxiety, stress, frustration, and depressive disorders (Kılıç et al., 2020). Hence, it has become vital to promote mechanisms that deal positively with mental health and tackle the social and mental effects of the pandemic (Matthews et al., 2018).

Broader research review from studies done in various countries such as Turkey, China, Germany, and USA, have displayed the influence of COVID-19 on population with respect to policies, healthcare, economy, etc. (Kılıç et al., 2020; Jungmann & Witthöft, 2020; Zhang & Ma, 2020; X. Zhang et al., 2020). A study from China conducted in January discussed long-term effects such as PTSD and the outcome of negative coping (Liang et al., 2020). A review of related works in India revealed that the primary focus had been on older adults, healthcare professionals, and at-risk patients (Dubey et al., 2020). This study focused on studying the effects of COVID-19 on the student population in India and gauge their response to knowledge, attitude, anxiety experience, and mental health care (Roy et al., 2020). It is imperative to assess the student population’s perspective, response, and progress during the pandemic, especially in a country like India where the majority of the population comprises of youth.

## Methods and material

Data collection, while usually done with personal contact and manually, proved to be challenge due to the lockdown and social distancing protocols for COVID-19. Thus, an online survey was designed considering the freedom of response, confidentiality and anonymity for the participants. Questions were framed based on basic demographic information, and perceptions about the impact of COVID-19 on-1) social impact (lockdown, strategies implemented, and general awareness), 2) personal impact (goal attainment, change in routine, financial stability and productivity, coping mechanisms, social media usage, relationships, etc.) and 3) psychological impact (stress, symptoms of anxiety & depression, suicidal ideation and addictive behaviour/drug indulgence) along with the perceptions about overall mental health. The responses were assessed on two scales: a five-point Likert scale for questions on personal impact (Strongly Agree, Agree, Partially Agree, Disagree and Strongly Disagree) and for questions on assessing the symptoms of impact on mental health a ten-point rating was used (10% during COVID 19 to 100% during COVID19). The data collection was kept open for a period of seven days and based on responses collected from the student participants of age 16-25 years. A total of N=351 responses were recorded. The data consisted mainly of students from the state of Maharashtra, which is worst affected by COVID-19 in India [2]. A detailed demographic description for the same is presented in Table 1.

**Table 1:**
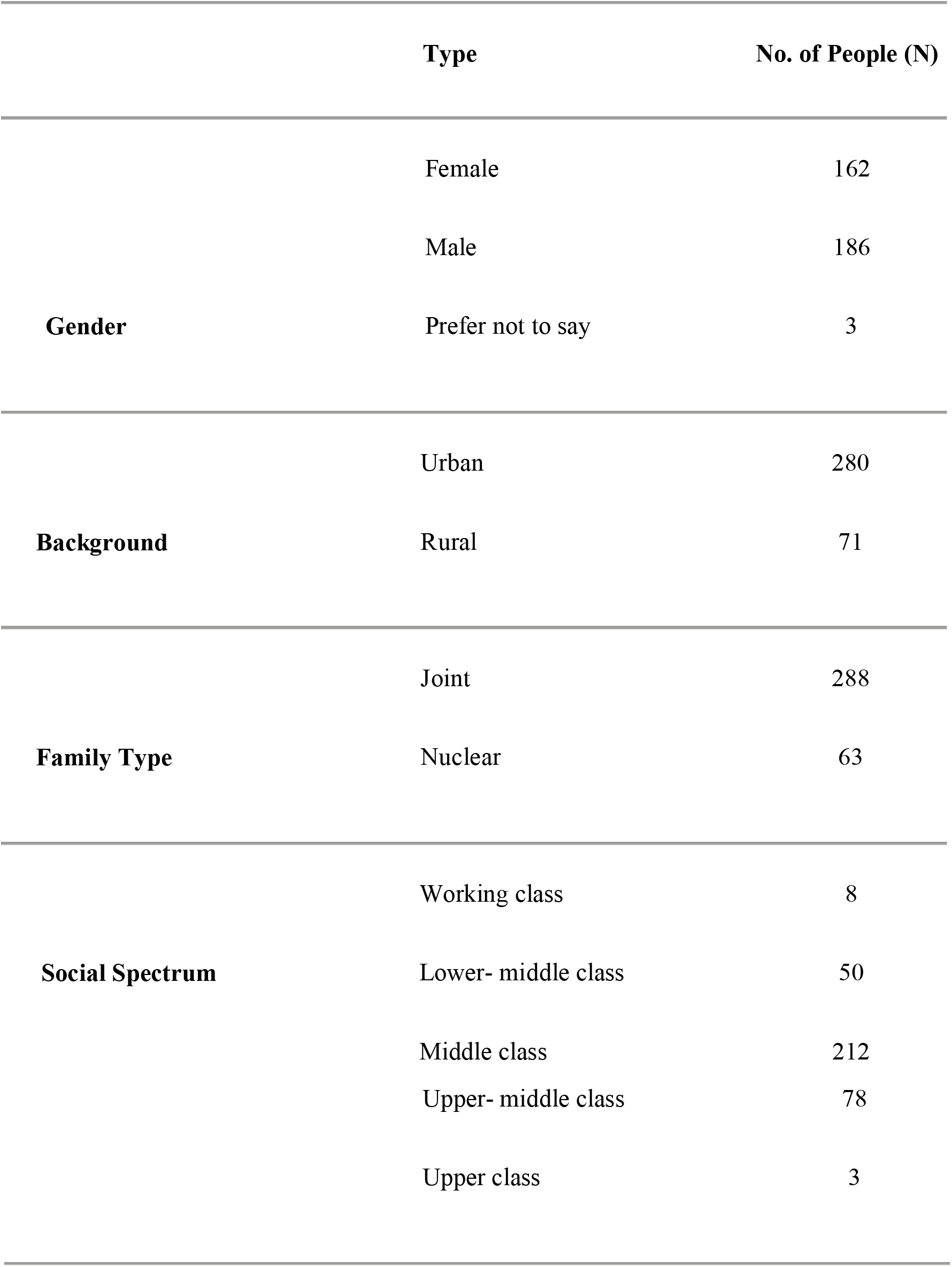
Demographic descriptions of N=351 student responses

For the analysis of the data obtained, Anaconda’s Jupyter Notebook was used. Also, Python’s Pandas, Seaborn libraries and SPSS. 20 were utilized for numerical analysis, visualizations and comparison of means, respectively. All the analysis was based on observing the trends of percentages, descriptive and parametric statistics (students t test) based on a systematic survey design. Since the data has been collected in the initial stage of the COVID 19 spread, it was limited to understanding the trend and not particularly to draw affirmative conclusions.

## Results

The results obtained are presented based on the analysis done for the social and psychological impact COVID 19 has made on the student sample

### 1. Social Impact

The first section of the survey assessed participants’ perspectives on the social scenario and its impact on them. Table 2 depicts that a majority of the respondents reported having sufficient information about the quarantine and following the protocols of social distancing. However, there is a variation in answer for Unlock 1.0 being introduced at the right time and COVID-19 eliminating social barriers. Further, participants reported an increase in awareness about mental health.

**Table 2:**
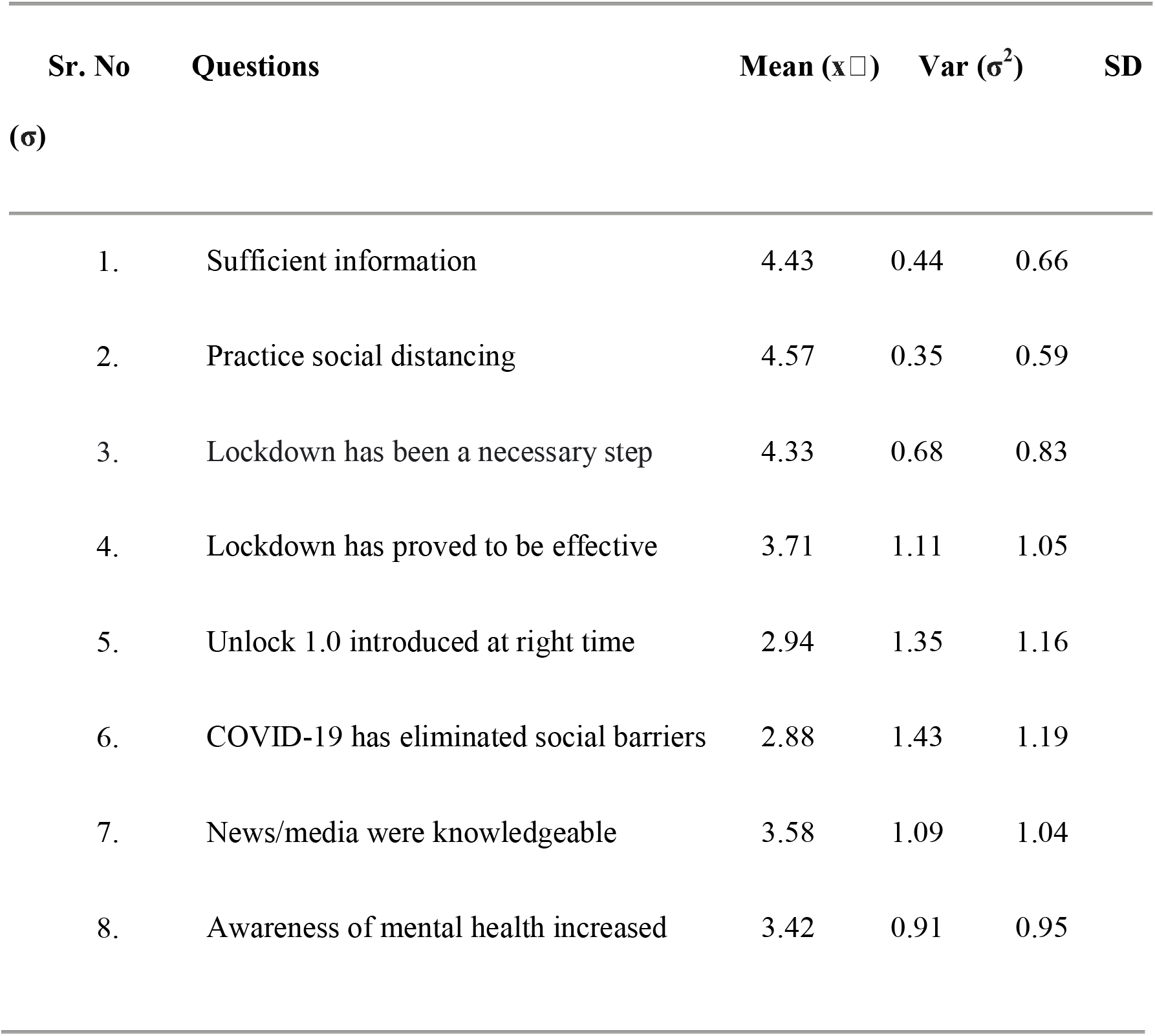
Response to Social Impact of COVID-19

| Sr. No | Questions | Mean ($\bar{x}$) | Var ($\sigma^2$) | SD |
| --- | --- | --- | --- | --- |
| ($\sigma$) | | | | |
| 1. | Sufficient information | 4.43 | 0.44 | 0.66 |
| 2. | Practice social distancing | 4.57 | 0.35 | 0.59 |
| 3. | Lockdown has been a necessary step | 4.33 | 0.68 | 0.83 |
| 4. | Lockdown has proved to be effective | 3.71 | 1.11 | 1.05 |
| 5. | Unlock 1.0 introduced at right time | 2.94 | 1.35 | 1.16 |
| 6. | COVID-19 has eliminated social barriers | 2.88 | 1.43 | 1.19 |
| 7. | News/media were knowledgeable | 3.58 | 1.09 | 1.04 |
| 8. | Awareness of mental health increased | 3.42 | 0.91 | 0.95 |

### 2. Personal Impact

The second part of the survey involved studying the students’ perspective on the impact COVID-19 has had on their personal lives. Table 3 shows that the majority of the participants have experienced adjustments in daily routine due to COVID-19. While the daily work regime has been affected negatively, an increase in the sleeping pattern has been observed. Furthermore, a rise in apprehension about productivity, postponing planned activities, and nervousness about healthcare availability was observed. Despite their attempts to create a new routine, an apprehension in everyday adjustments seem evident from the responses.

**Table 3:**
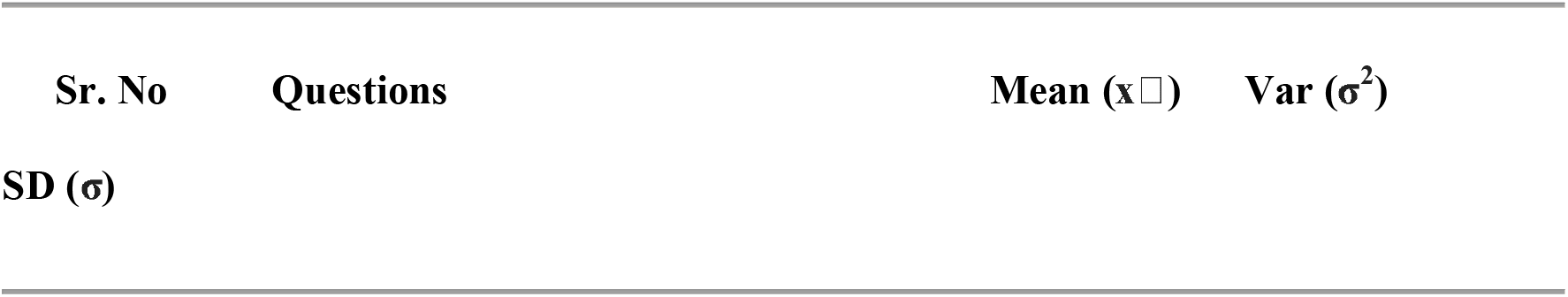
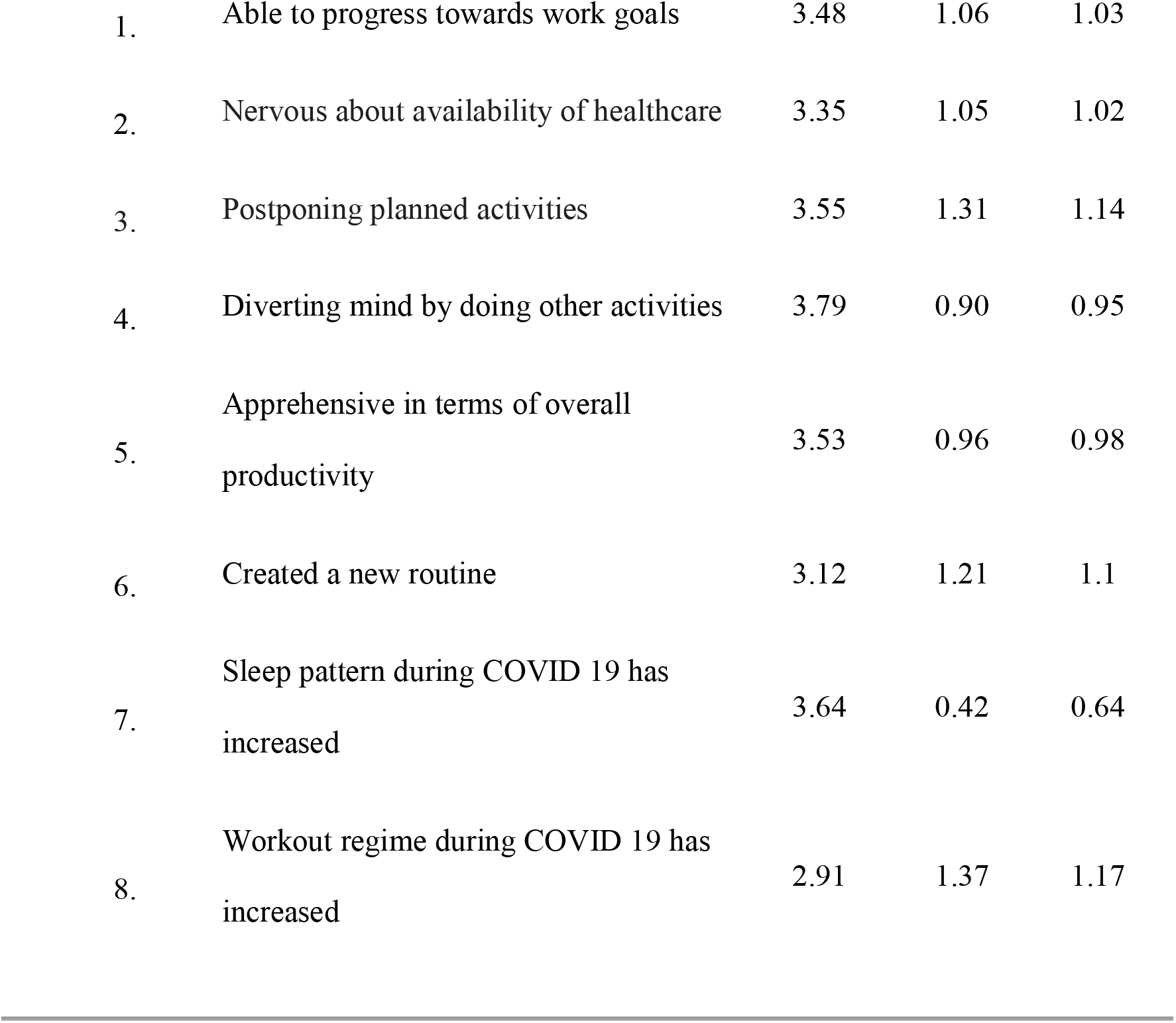
Response to Personal Impact of COVID-19

| Sr. No | Questions | Mean ($\bar{x}$) | Var ($\sigma^2$) |
| --- | --- | --- | --- |
| SD ($\sigma$) | | | |

|  |  |  |  |  |
| --- | --- | --- | --- | --- |
| 1. | Able to progress towards work goals | 3.48 | 1.06 | 1.03 |
| 2. | Nervous about availability of healthcare | 3.35 | 1.05 | 1.02 |
| 3. | Postponing planned activities | 3.55 | 1.31 | 1.14 |
| 4. | Diverting mind by doing other activities | 3.79 | 0.90 | 0.95 |
| 5. | Apprehensive in terms of overall productivity | 3.53 | 0.96 | 0.98 |
| 6. | Created a new routine | 3.12 | 1.21 | 1.1 |
| 7. | Sleep pattern during COVID 19 has increased | 3.64 | 0.42 | 0.64 |
| 8. | Workout regime during COVID 19 has increased | 2.91 | 1.37 | 1.17 |

### 3. Psychological Impact

Table 4 demonstrates the evaluation of response to perceptions about the psychological impact of COVID-19 on participants. An increase in overall symptoms of stress, anxiety and an overall increase in physiological symptoms is observed; although not very high this is not insignificant. Further, both females and male students responded to feeling the symptoms more than before or to a higher extent post outbreak. While females are plagued with symptoms of over thinking and random thoughts, males are feeling more socially withdrawn or feelings of self-harm.

**Table 4:**
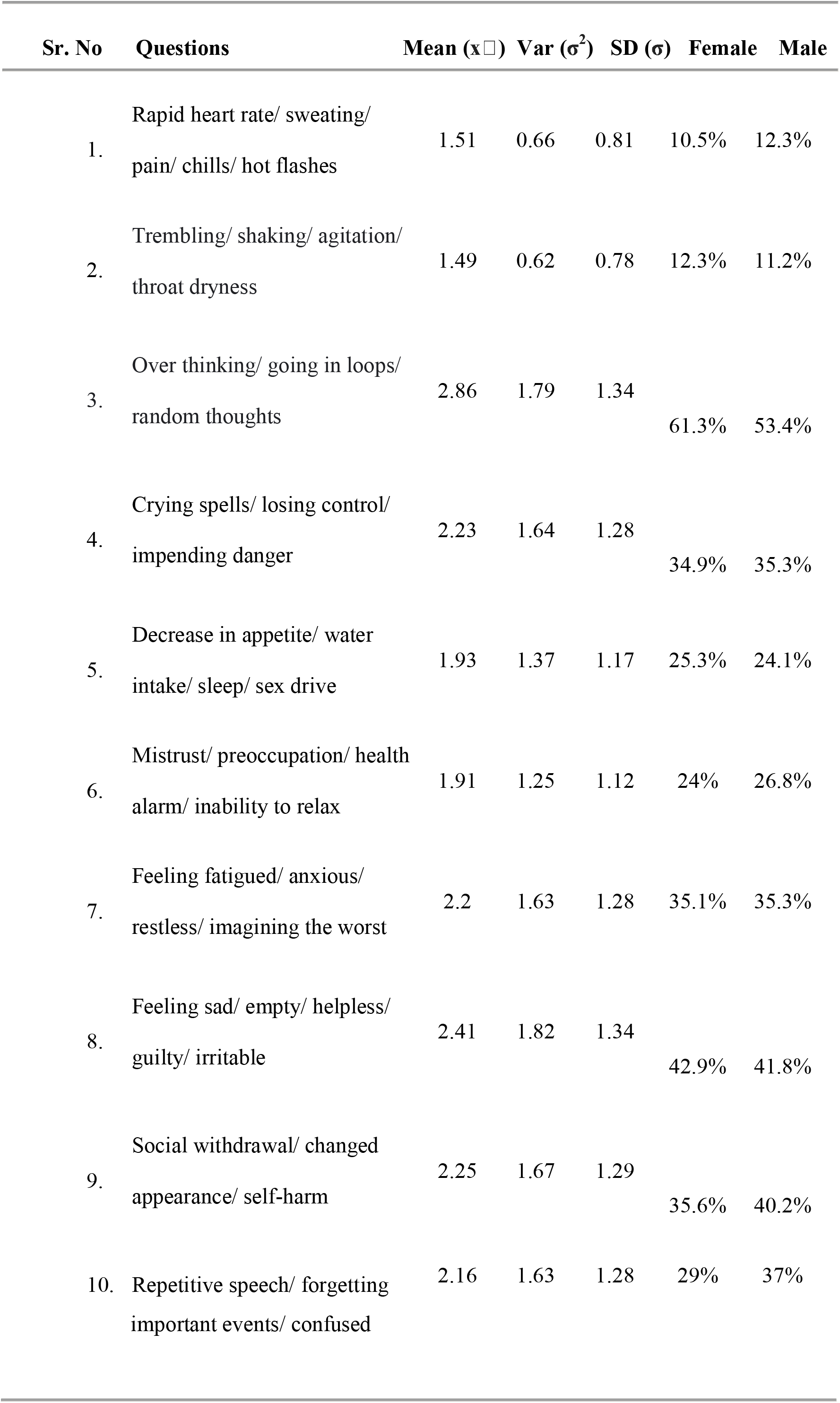
Response to Psychological Impact of COVID-19 and comparison based on gender

The data was also analysed by comparing the differences between the responses as per the demographics. A comparison between male and female responses for a set of psychological questions is covered in the form of a bar plot in Figure 1(a), while Figure 1(b) shows comparative analysis based on rural and urban background for responses. Female students’ mean responses for all questions are higher than their counterparts except for indulgence in intoxicants reported as low by both. As for urban and rural students, mean scores for urban background students are more than students in rural areas; both show an increase in online activities, while indulgence in intoxicants is reported to be lowest. The descriptive statistics for these groups are depicted in Table 5.

**Table 5:** Descriptive statistics of COVID-19 response for the sample based on Gender and Background

| Groups/Parameters |  | Time on social media |  |  | Apprehensive about career |  |  | Positively close to family |  |  | Conscious about diet and food intake |  |  |
| --- | --- | --- | --- | --- | --- | --- | --- | --- | --- | --- | --- | --- | --- |
|  |  | Mean | Var | SD | Mean | Var | SD | Mean | Var | SD | Mean | Var | SD |
| | | ($\bar{x}$) | ($\sigma^2$) | ($\sigma$) | ($\bar{x}$) | ($\sigma^2$) | ($\sigma$) | ($\bar{x}$) | ($\sigma^2$) | ($\sigma$) | ($\bar{x}$) | ($\sigma^2$) | ($\sigma$) |
| Gender | Female | 3.66 | 1.59 | 1.26 | 3.78 | 1.26 | 1.12 | 3.85 | 0.99 | 0.99 | 3.66 | 0.7 | 0.84 |
|  | Male | 3.82 | 1.46 | 1.21 | 3.51 | 1.39 | 1.17 | 3.64 | 1.11 | 1.05 | 3.59 | 0.97 | 0.58 |
| Background | Rural | 4.02 | 1.35 | 1.16 | 3.64 | 1.16 | 1.07 | 3.79 | 1.04 | 1.02 | 3.61 | 0.86 | 0.92 |
|  | Urban | 3.67 | 1.55 | 1.24 | 3.65 | 1.39 | 1.18 | 3.72 | 1.08 | 1.04 | 3.63 | 0.86 | 0.92 |
| Total |  | 3.75 | 1.52 | 1.23 | 3.65 | 0.96 | 1.16 | 3.73 | 1.07 | 1.03 | 3.62 | 0.85 | 0.92 |

n (males)= 186, n (females)= 162; n (urban)= 280, n (rural)= 71

**Figure 1:**
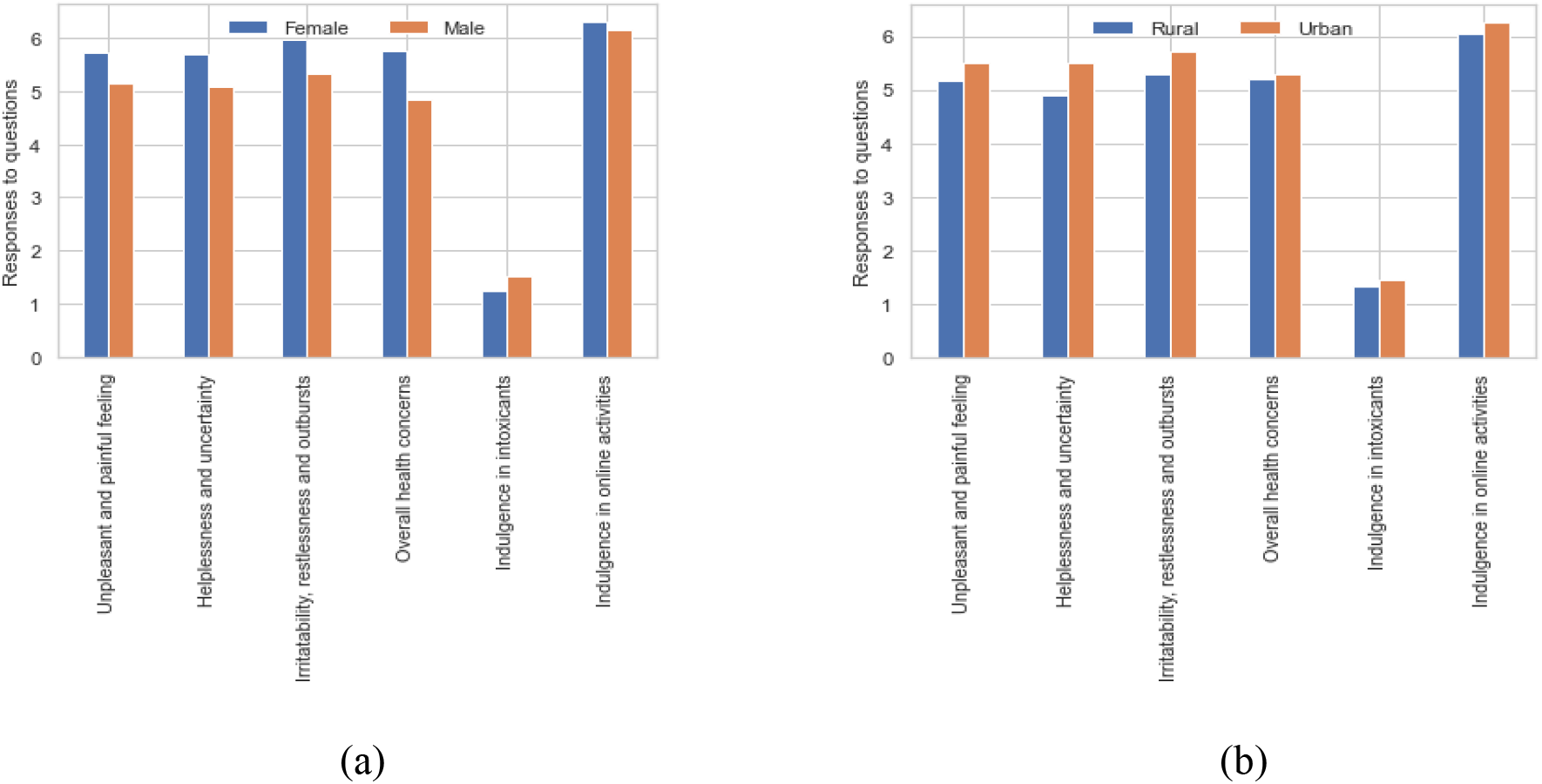
Bar graph representation of comparative statistic for response to questions, **(a)** Based on Gender: Female/Male; **(b)** Based on Background: Rural/Urban.

To find significant evidence for the expected differences between the groups, students t test (equal variance assumed) was done using SPSS and a comparative effect of COVID-19 based on gender and background was calculated, as represented in Tables 6 for social and personal impact and psychological impact. The level of significance was set at p≤0.05.

**Table 6:**
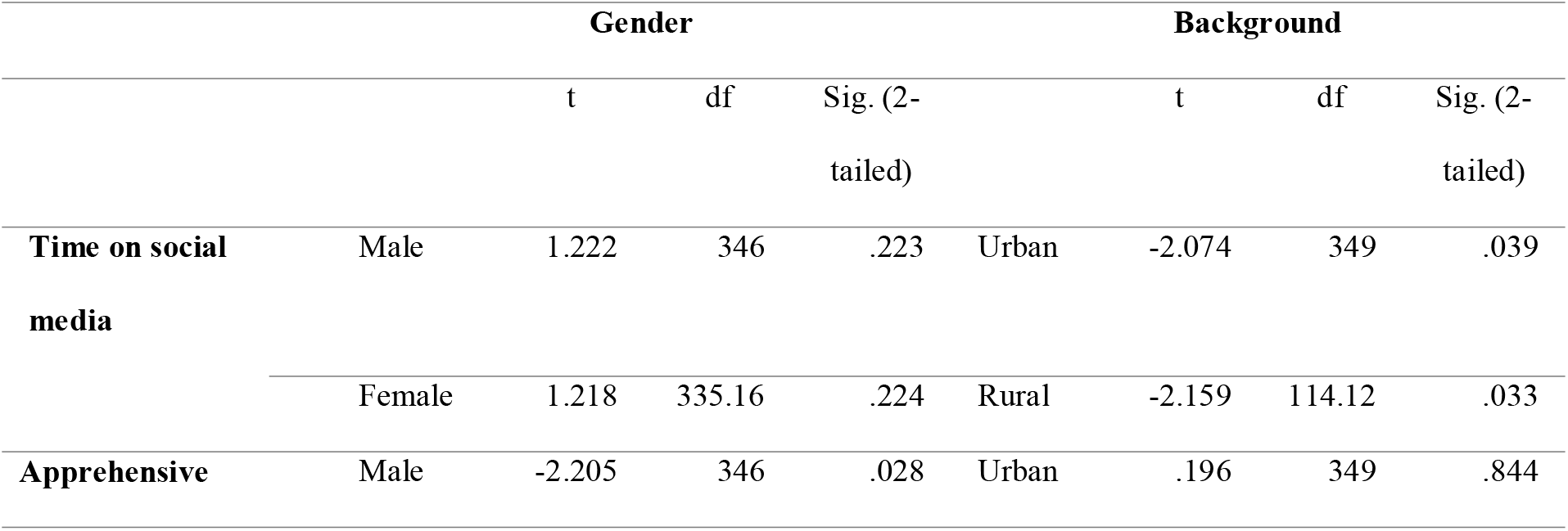
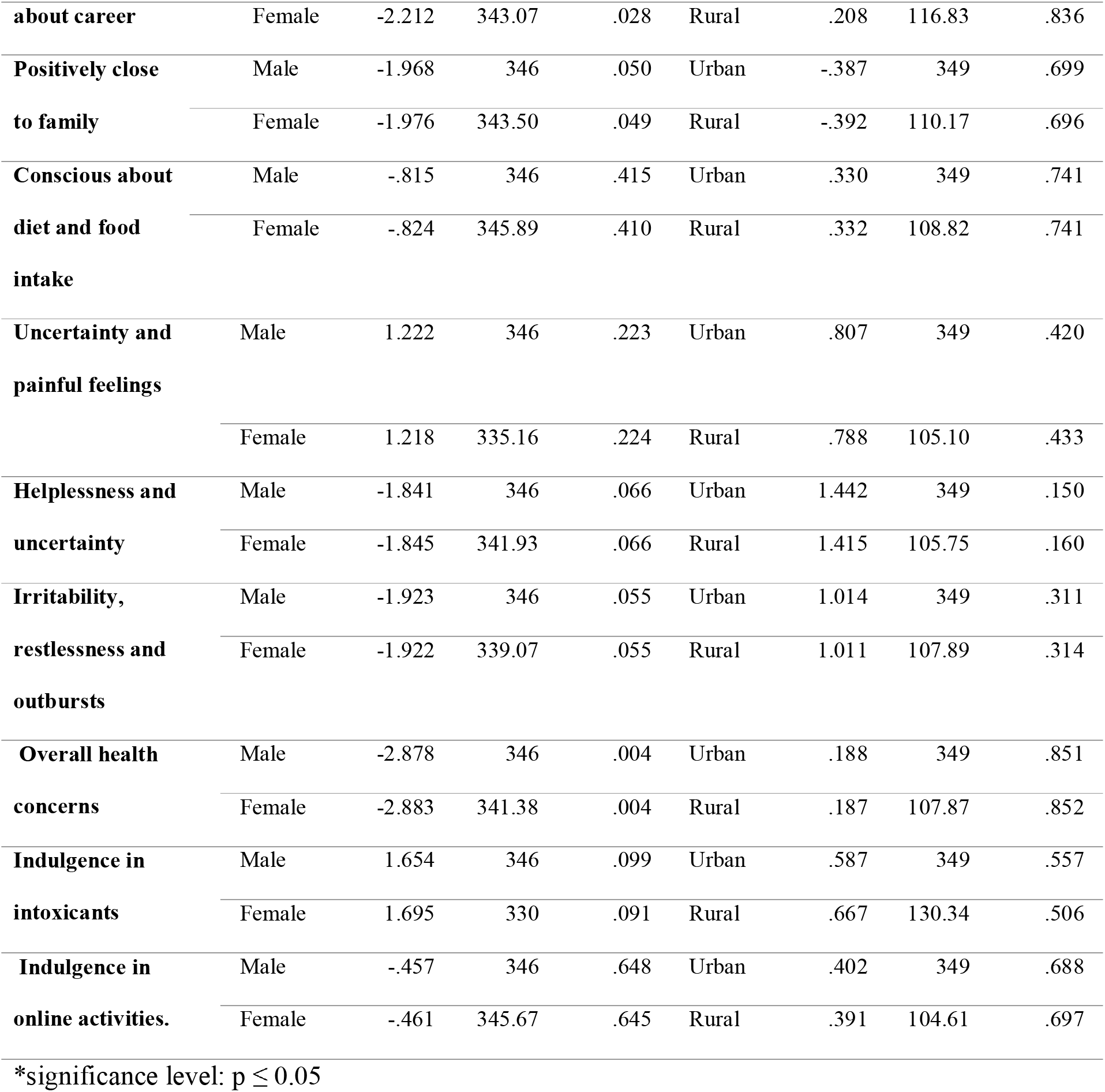
Comparison of independent samples’ mean of COVID-19 response based on Gender and Background: Social & Personal impact

It was found that females (M= 3.78, SD= 1.17) were significantly more apprehensive about career and future prospects than males (M= 3.51, SD= 1.12), and also females (M= 3.85, SD= 0.99) felt significantly positively close to the family during lockdown as compared to males (M= 3.63, SD= 1.05). As far as background was concerned, students from rural background (M= 4.01, SD= 1.16) significantly spent more time on social media as compared to those from urban background (M= 3.68, SD= 1.25). Further, in the psychological impact while there was significant impact on the levels of irritability, restlessness and outburst, a noticeable difference was found in the overall health concerns where females (M= 5.77, SD= 2.93) appeared to be more affected than males (M= 4.85, SD= 2.99). No significant difference between the groups was found on other factors of social, personal and psychological impact.

While COVID19 has affected people on various levels-social, personal, psychological, physical, etc.-an increase in inclination towards existential approach and introspection can be generally seen. Hence, to understand students’ perspective, responses show a high inclination being more practical, realistic and prepared, followed by becoming self-aware and enjoying solitude (Table 7). The youth of this country appears to be very pragmatic while working on solitude.

**Table 7:**
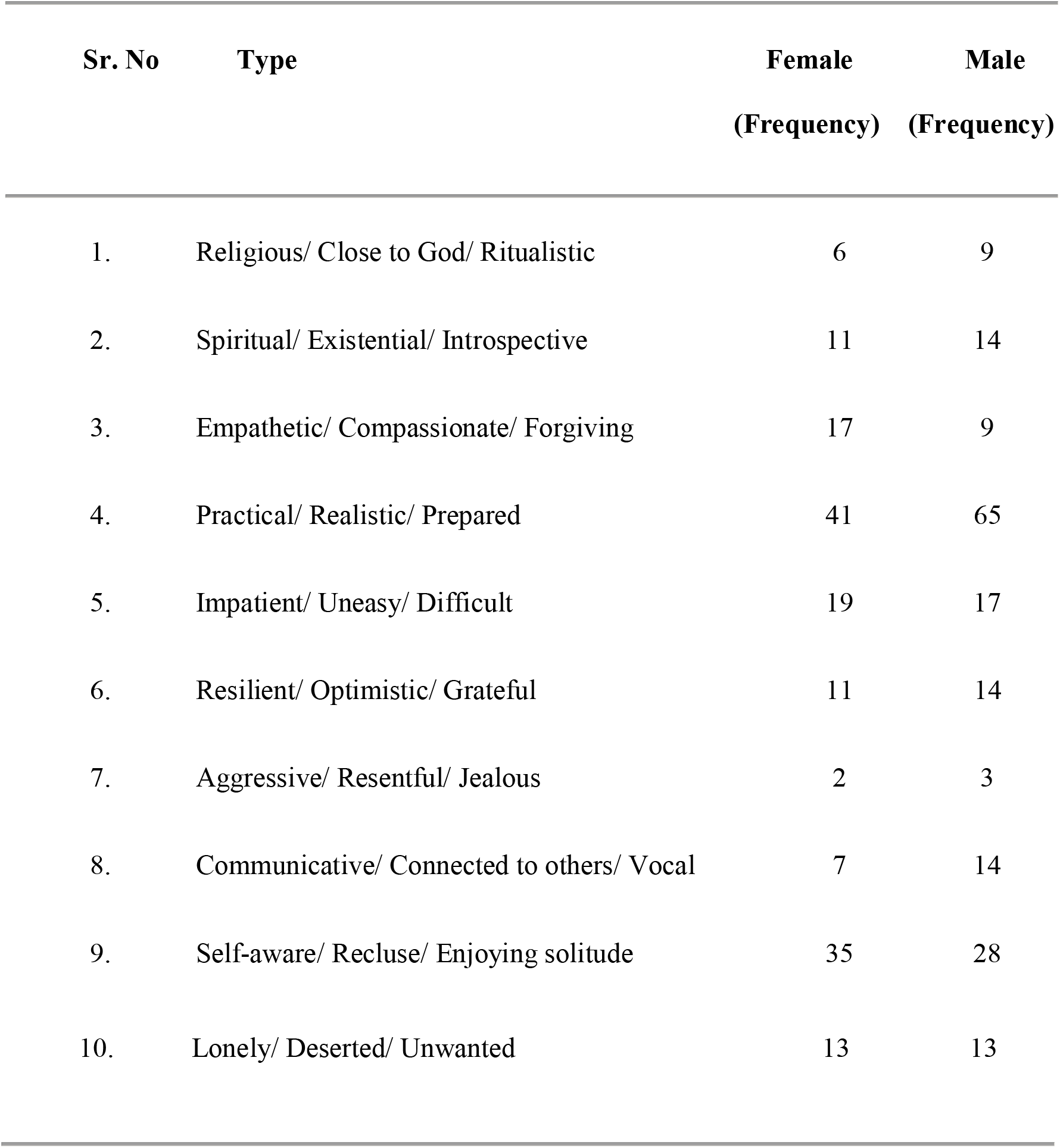
Comparison of COVID-19: Effect on perceptions about self/life based on gender

Further, reaching out to a mental health professional is more acceptable to females (70%), urban students (65% and those from nuclear family (63%), while 48% rural students, 55% males and 56% students belonging to joint families indicate a low inclination, as shown in Fig 2(a). For the student population, a significant impact of COVID19 has been on the reduced proximity among family and friends. Figure 2(b) shows the number of students affected positively while living with family. Rural students (61%) and females (65%) perceive staying with family as positive unlike their counterparts.

**Figure 2:**
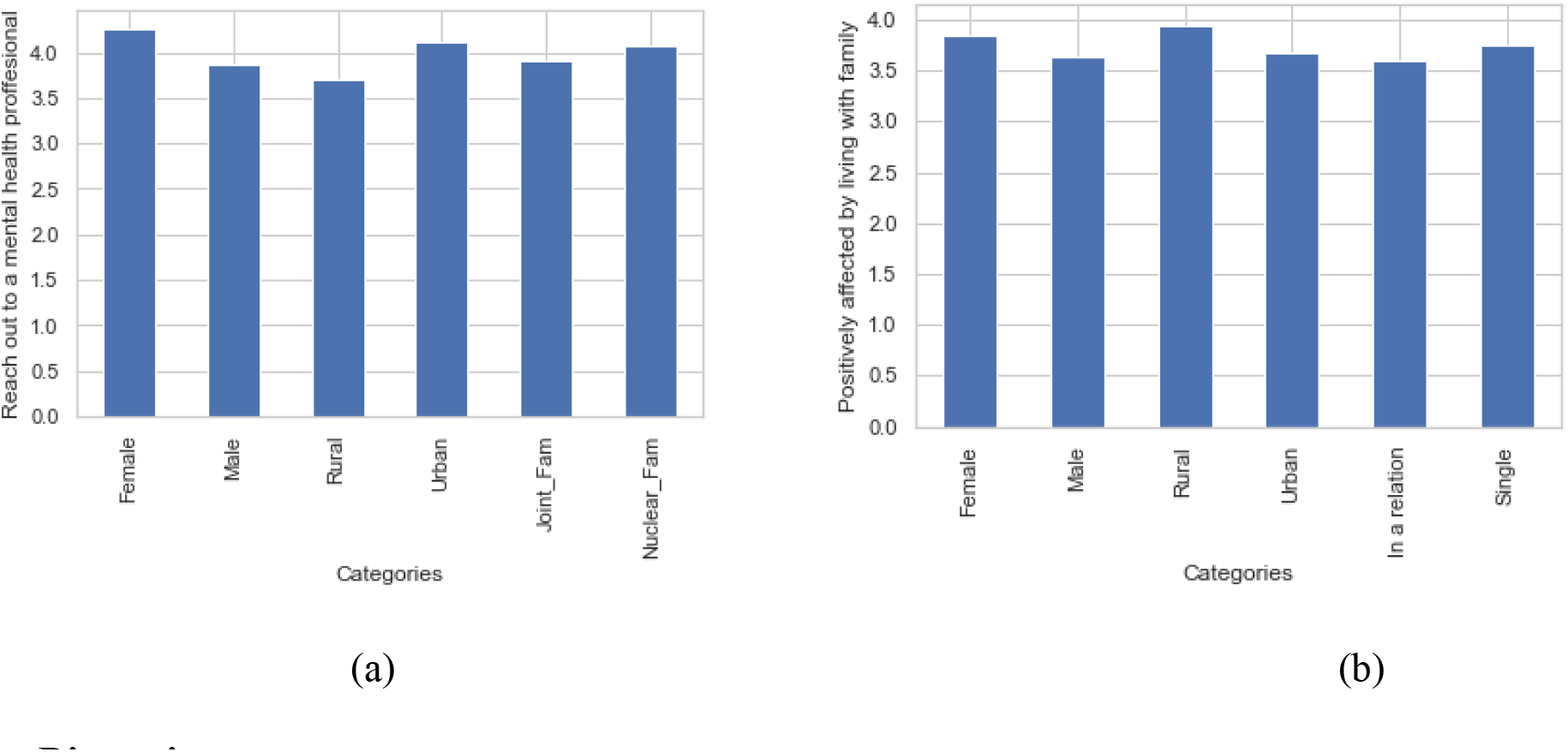
Bar graph representation of comparative statistic, **(a)** No. of people that will reach out to mental health professional; **(b)** No. of people reporting being positively affected by living with family.

## Discussion

Generally, the student population seem to be equipped with the basic information, aware of the precautions, measures and abided by the lockdown rules. However, the popular belief that the outbreak has eliminated social barriers and made everyone equal is received with mixed responses. This can be attributed to the fact that financial or health risks continue to increase and while information is sufficient, opportunities or facilities may not be. There is also a divide in the opinion regarding Unlock 1.0 introduced in India on 1 June, especially since the cases continue to rise exponentially. The news and media reports may have been an influencing factor in these responses.

The pandemic has also proven to be a test of adjustment in a novel situation never experienced before. While the workout regime seems to have taken a leap, the data seem to be spread-out. This indicates that while it is easy to accept that regular workout and sleep patterns have been affected, it is seemingly difficult to accept that the same has happened with productivity. From the general trend it can be seen that the students in particular are not clinically affected. However, over thinking and apprehensions about the future seem to be affecting the females more than the males, which is noticeable since in a society like India, the males are considered to be the primary earning members of a family. This indicates a shift in the thought process among the youth of this country, which is certainly positive. Also, females seem to be more health conscious and feel more positive about being close to family during this crisis as compared to males. Although this difference in perception about social support is clear among males and females, feelings of sadness and emptiness are similar and symptoms of depression seem to be affecting the youth equally, irrespective of the gender. Hence, males seem to be turning more withdrawn, probably as part of coping, and women towards building stronger bonds with family. Also, an overall increase in irritability, overall health concerns and feeling unpleasant is seen more in females than males; the indulgence in drugs and other intoxicants is however the opposite with males scoring over females, although the difference is not much (Smojver-Ažić & Bezinović, 2011; Wardle et al., 2004). The males also seem to have become more prone to solitude, desired isolation and perceived self-harm which is not only indicative of suicidal thoughts but is alarming as well. This suggests the impact of increased ambiguity due to COVID-19 leading to helplessness. Students reporting a change in personality and feelings of self-harm cannot be neglected and should be provided with appropriate intervention measures. Therefore, a gender specific approach in trying to overcome effects of pandemic is also important.

When it comes to students coping with COVID-19, the urban student scores in unpleasant feelings, irritability, helplessness and uncertainty with a noticeable difference. This might mean that though used to the fast-paced lifestyle and corporate culture, the urban youth finds a crisis hard to handle due to the societal pressure and demands being higher in cities than in rural areas. According to research on mental health in large cities, the risk for some major mental illnesses (e.g. anxiety, psychotic, mood, or addictive disorders) is generally higher (Gruebner et al., 2017).

The results on internet usage by urban and rural students is almost the same, which indicates the wide reach of online facilities into the remote areas of the country; a developing nation such as India seems to be advancing towards technological growth not just in its cities but also with its backbone-the rural India.

With the unanimous response of increased awareness about mental health, the responses to reaching out to a mental health professional vary. Rural students seem to be less agreeable to reach out to a mental health professional as compared to the urban population, which may be attributed to the lack of awareness about mental health. The reasons could be the social dependence, fear of labelling and stigma affect the identification, treatment and maintenance of mental health problems (Nicholson, 2008). Further, rural students tend to be positive about being close to family since they usually live closer to nature dominant life, as opposed to the urban students. Hence, they have a great bond of relationship and good affinity (Marisennayya, 2020).

While the overall impact on mental health during the pandemic is not as high as suspected, a social aspect is worth mentioning. Majority of students n=165) accounts “living with near and dear ones” as a primary reason for affecting mental health as compared to “living away from near and dear ones” (n=64) among other factors such as online activities, imposed restrictions, new updates, etc. This is interesting, since the overall results are indicative of less impact on mental health, primarily due to the availability of near and dear ones. While India is a collectivistic society that ensures the psychological dependence on family and friends, the student population seem to be deviating from the norm. It would be interesting to study the differences regarding this between students and the working adults of the country. Overall, while the current paradigm is unexpected and has created a state of panic around the world, its effect on students’ mental health in India compared to western countries is lesser. Even though social distancing and isolation has changed dynamics of friendships and relationships making it virtual, being close to family and loved ones has acted as a support system, albeit with reservation. By and large, a change in perspective has been observed regarding mental health, which can certainly be rendered helpful in adjusting with the new normal.

This study was conducted keeping in mind the student population of the most affected state in India by the global pandemic COVID-19 and the sample used here is representative of this population only. There will certainly be differences in responses if this study is conducted in other parts of the country or otherwise. The small sample size and by extension the limited methods used for statistical analysis are the limitations of this study. A replication of the present research, if done nation-wide, would provide a more representative data for generalization.

## Conclusion

The current situation of COVID-19 has affected the students’ mental health, specifically in some aspects. Although students are finding out ways to deal with the uncertain situation for example, creating a schedule for daily activities, getting involved in developing a skill, increasing use of social media for entertainment and also to gain information about safety measures, the repercussions of uncertainty, feelings of depression and differences in male-female and urban-rural students can be seen on various parameters. The increasing practicality and independence among the youth in a way questions the interdependent nature of the society. However, the role and importance of family and friends cannot be undermined and remains a crucial factor in handling crises. An increase in mental health awareness, gender-based intervention strategies, relevant coping mechanisms for the youth from varied backgrounds may be devised to help the students cope with the issues related to mental health, during a pandemic, or otherwise.

## Limitations & Further Scope

This study was conducted keeping in mind the student population of the most affected state in India by the global pandemic COVID-19 and the sample used here is representative of this population only. There may certainly be differences in responses if this study is conducted in other parts of the country or otherwise. The small sample size and by extension the limited methods used for statistical analysis are the limitations of this study. A replication of the present research, if done nation-wide, would provide a more representative data for generalization.

## Data Availability

The data was collected from the consenting participants via online survey.

## Acknowledgements

The authors acknowledge the constant support of the Director and Deputy Director, and the Department Head of Applied Sciences, College of Engineering, Pune and are thankful for the guidance provided throughout this project.

## References

Coronavirus Disease (COVID-19) - events as they happen. (n.d.). Retrieved from https://www.who.int/emergencies/diseases/novel-coronavirus-2019/events-as-they-happen

Coronavirus Disease (COVID-19) Situation Reports. (n.d.). Retrieved from https://www.who.int/emergencies/diseases/novel-coronavirus-2019/situation-reports/

Dubey, S., Biswas, P., Ghosh, R., Chatterjee, S., Dubey, M. J., Chatterjee, S., Lahiri, D., & Lavie, C. J. (2020). Psychosocial impact of COVID-19. Diabetes & Metabolic Syndrome: Clinical Research & Reviews, 14(5), 779–788. https://doi.org/10.1016/j.dsx.2020.05.035

Ebrahim, S. H., Ahmed, Q. A., Gozzer, E., Schlagenhauf, P., & Memish, Z. A. (2020). Covid-19 and community mitigation strategies in a pandemic. BMJ, m1066. https://doi.org/10.1136/bmj.m1066

Fiorillo, A., & Gorwood, P. (2020). The consequences of the COVID-19 pandemic on mental health and implications for clinical practice. European Psychiatry, 63(1). https://doi.org/10.1192/j.eurpsy.2020.35

Gruebner, O., Rapp, M. A., Adli, M., Kluge, U., Galea, S., & Heinz, A. (2017). Cities and Mental Health. Deutsches Aerzteblatt Online. https://doi.org/10.3238/arztebl.2017.0121

He, G., Sun, W., Fang, P., Huang, J., Gamber, M., Cai, J., & Wu, J. (2020). The clinical feature of silent infections of novel coronavirus infection (COVIDC19) in Wenzhou. Journal of Medical Virology, 92(10), 1761–1763. https://doi.org/10.1002/jmv.25861

Jungmann, S. M., & Witthöft, M. (2020). Health anxiety, cyberchondria, and coping in the current COVID-19 pandemic: Which factors are related to coronavirus anxiety? Journal of Anxiety Disorders, 73, 102239. https://doi.org/10.1016/j.janxdis.2020.102239

Kiliç, T. ş. i., Bostan, S., Erdem, R., Öztürk, Y. E., & Yılmaz, A. (2020). The Effect of COVID-19 Pandemic on the Turkish Society. Electronic Journal of General Medicine, 17(6), em237. https://doi.org/10.29333/ejgm/7944

Liang, L., Ren, H., Cao, R., Hu, Y., Qin, Z., Li, C., & Mei, S. (2020). The Effect of COVID-19 on Youth Mental Health. Psychiatric Quarterly, 91(3), 841–852. https://doi.org/10.1007/s11126-020-09744-3

Mahmoud, J. S. R., Staten, R. “. T. “., Hall, L. A., & Lennie, T. A. (2012). The Relationship among Young Adult College Students’ Depression, Anxiety, Stress, Demographics, Life Satisfaction, and Coping Styles. Issues in Mental Health Nursing, 33(3), 149–156. https://doi.org/10.3109/01612840.2011.632708

Marisennayya, Senapathy. (2020). Re: What are the characteristics of rural and urban development? What are the differences and similarities?. Retrieved from: https://www.researchgate.net/post/What_are_the_characteristics_of_rural_and_urban_development_What_are_the_differences_and_similarities/5e5a67b3f0fb629c793fbdac/citation/download

Matthews, T., Danese, A., Caspi, A., Fisher, H. L., Goldman-Mellor, S., Kepa, A., Moffitt, T. E., Odgers, C. L., & Arseneault, L. (2018). Lonely young adults in modern Britain: findings from an epidemiological cohort study. Psychological Medicine, 49(2), 268–277. https://doi.org/10.1017/s0033291718000788

Nicholson, L. A. (2008). Rural mental health. Advances in Psychiatric Treatment, 14(4), 302–311. https://doi.org/10.1192/apt.bp.107.005009

Pneumonia of unknown cause – China. (2020, January 30). Retrieved from https://www.who.int/csr/don/05-january-2020-pneumonia-of-unkown-cause-china/en/

Roy, D., Tripathy, S., Kar, S. K., Sharma, N., Verma, S. K., & Kaushal, V. (2020). Study of knowledge, attitude, anxiety & perceived mental healthcare need in Indian population during COVID-19 pandemic. Asian Journal of Psychiatry, 51, 102083. https://doi.org/10.1016/j.ajp.2020.102083

Sharma, V. K., & Nigam, U. (2020). Modeling and Forecasting of Covid-19 Growth Curve in India. MedRxiv. https://doi.org/10.1101/2020.05.20.20107540

Smojver-Ažić, S., & Bezinović, P. (2011). Sex differences in patterns of relations between family interactions and depressive symptoms in adolescents. Croatian Medical Journal, 52(4), 469–477. https://doi.org/10.3325/cmj.2011.52.469

Wardle, J., Haase, A. M., Steptoe, A., Nillapun, M., Jonwutiwes, K., & Bellisie, F. (2004). Gender differences in food choice: The contribution of health beliefs and dieting. Annals of Behavioral Medicine, 27(2), 107–116. https://doi.org/10.1207/s15324796abm2702_5

Zhang, X., Wang, Y., Lyu, H., Zhang, Y., Liu, Y., & Luo, J. (2020). The Influence of COVID-19 on Well-Being. https://doi.org/10.31234/osf.io/znj7h

Zhang, Y., & Ma, Z. F. (2020). Impact of the COVID-19 Pandemic on Mental Health and Quality of Life among Local Residents in Liaoning Province, China: A Cross-Sectional Study. International Journal of Environmental Research and Public Health, 17(7), 2381. https://doi.org/10.3390/ijerph17072381

